# Cognitively healthy young adults with *APOEe4* gene show disrupted functional connectivity of graph properties in multiple resting-state networks

**DOI:** 10.1101/2024.09.27.24314481

**Authors:** Ludmila Kucikova, Jianmin Zeng, Adam J. Brass, Carlos Muñoz-Neira, Craig W. Ritchie, Graciela Muniz-Terrera, John O’Brien, Li Su

**Author notes:** Corresponding author: Li Su; Neuroscience Institute, Medical School, University of Sheffield, Sheffield, S10 2HQ, UK; Department of Psychiatry, School of Clinical Medicine, University of Cambridge, Cambridge, CB2 0SZ, UK. These authors contributed equally (Ludmila Kucikova, Jianmin Zeng).

## Abstract

The *apolipoprotein (APOE) e4* allele is associated with brain changes in healthy carriers that are similar to changes observed in patients with Alzheimer’s Disease, including abnormalities in functional connectivity. The trajectory of these changes across the lifespan, specifically in early adulthood is still not clear. This study explores the link between the *APOE* genotype and functional connectivity in 129 cognitively healthy Chinese Han college students (aged 17-22 years). By using graph theory, we assessed the connectivity in seven resting-state networks of interest using three different thresholding methods and three different forms of network parcellation. Average Path Length and Closeness Centrality were disrupted in *e4* carriers in the sensorimotor, visual, salience, and Default Mode salience networks; with effects replicated using different thresholding but not different parcellation methods. This study demonstrated the genetics-related vulnerability in the brain of young *APOEe4* carriers across multiple resting-state networks.

## 1. Introduction

The *apolipoprotein e4* (*APOEe4*) allele is a strong genetic risk factor for Alzheimer’s Disease (AD)^1^. Despite being widely recognised for its role in lipid transport^2^, amyloid beta clearance^3^, and neurofibrillary tangle formation^4^, the impact of *APOEe4* on brain function and structure might begin long before AD-related pathology occurs.

Functional brain imaging studies have shown altered brain function in *APOEe4* carriers already before the onset of clinical symptoms, specifically in the activity and connectivity of resting-state networks in middle-aged individuals (reviewed^5,6^). A decrease in functional connectivity in the areas of the Default Mode Network (DMN) has been consistently reported in middle-aged adults. The findings in young adults are much more mixed with studies reporting both an increase^7–9^ and a decrease^9–13^. Only the medial visual network^13^ showed alterations in young *APOEe4* carriers beyond the findings from the DMN. There were no differences in other resting-state networks including the executive/frontoparietal, visual, sensorimotor^10,13^, dorsoattentional, salience, and language^10^ and auditory networks^13^.

The inconsistencies in findings might be partly explained by the ethnicity of the participants^14^. While ethnicity is not consistently reported across all the studies, studies that were conducted on the Chinese Han population showed similarities in the directionality of connectivity differences. The connectivity decreased in the angular gyrus^10^, dorsolateral prefrontal cortex^12^, and between the hippocampus and praecuneus/posterior cingulate cortex and subgenual anterior cingulate cortex^9^. Moreover, recent follow-up research shows that the whole-brain connectivity in the young Chinese Han population decreases over time^15^. Additionally, evidence from other neuroimaging studies suggests that the presence of the *APOEe4* allele modulates brain structure and function in this population in young adulthood beyond connectivity^16,17^.

Further, the inconsistencies in the directionality observed across young adulthood might relate to subtle differences in brain organisation of resting-state networks in early adulthood. Therefore, using the measures that are more sensitive to subtle nuances and offer a more holistic view of brain alterations might offer deeper insights. Graph theoretical analysis has become an important tool in the examination of brain organisation and reorganisation at various stages of neurological and psychiatric disorders^18^. This approach enables a more comprehensive understanding of the intricate relationship of dynamic changes within the neural circuits associated with different stages of neurological and psychiatric conditions.

In neurodegeneration, studies suggest that graph properties undergo progressive alterations in Mild Cognitive Impairment (MCI) and AD^19^. These alterations are also evident in the functional networks of *APOEe4* carriers experiencing subjective cognitive decline^20^. Moreover, by using the combination of graph theory and machine learning approaches, Hojjati and colleagues^21^ identified areas underlying conversion from MCI to AD.

While some evidence exists from the cognitively healthy older population at risk of AD^22^, further exploration in pre-symptomatic stages remains limited with a notable gap in the literature concerning graph theoretical studies of functional networks in young adults (reviewed^23^). It is unclear what exact connectivity differences occur in at-risk populations and how early they begin. The lack of evidence emphasises the necessity for systematic exploration in this critical demographic. Investigating early disease vulnerabilities and monitoring disease progression across the lifespan is critical to understanding the nature of the disorder fully and to developing potential therapeutic and preventative interventions that might include risk factor modification.

In this work, we used a graph theoretical approach to investigate the behaviour of the resting-state networks in young *APOEe4* carriers. We analysed seven networks of interest: the DMN, sensorimotor, visual, salience, dorsoattentional, language, and frontoparietal networks. By exploring these networks, we aimed to unveil the pattern of connectivity alterations associated with genetic vulnerability in the brain. We were interested in the properties that relate to network communication, information flow, and organisation; namely, Betweenness Centrality, Closeness Centrality, Degree Centrality, Average Path Length, Global Efficiency, and Clustering Coefficient.

## 2. Methods

### 2.1. Study sample

A total of 392 young cognitively healthy self-declared Chinese Han college students from Southwest University participated in this study as an extension of the PREVENT-Dementia study^24^. The study followed the regulations of the PREVENT-Dementia and Southwest University. Research ethics was provided by Southwest University’s local ethics committee. All participants provided informed written consent.

All participants provided a saliva sample for DNA genotyping, which was determined by the Mass Array system (Agena iPLEX assay, San Diego, USA). For the purpose of this study, the participants were classified as either carriers (i.e., at least one copy of the *APOEe4* allele) or non-carriers (i.e., no copy of the *APOEe4* allele). A subset of 155 participants underwent neuroimaging using MRI. After excluding data from participants with poor segmentation or acquisition quality, a final dataset resulted in 129 participants. In total, there were 27 carriers (female = 16) and 102 non-carriers (female = 54). There were no significant differences between the gender of carriers (59% female) and non-carriers (53% female), age of carriers (19.6±0/98) and non-carriers (19.6±0.9), and years of education of carriers (12.9±0.55) and non-carriers (13.1±0.59).

### 2.2. Neuroimaging acquisition and pre-processing

MRI imaging data were acquired on a 3T Siemens whole-body scanner at Southwest University. Participants were instructed to keep their eyes closed and not to think about anything specific while they were scanned at rest. The study protocol and pre-processing are identical to our previously published work^10^.

In brief, resting-state echo planar images and T1-weighted MPRAGE anatomical images were obtained. Neuroimaging pre-processing and functional connectivity analysis were performed using the CONN software (https://www.nitrc.org/projects/conn, RRID:SCR_009550) and in-house MatLab scripts (MathWorks, Natrick, MA). The pre-processing sequence in the CONN included motion estimation and correction, correction for inter-slice differences in acquisition time, outlier detection, segmentation, MNI152 co-registration, and smoothing with a Gaussian kernel of 8mm full-width half maximum. In addition, functional data were denoised by regressing potential confounding effects characterised by white matter timeseries, cerebrospinal fluid timeseries, motion parameters and their first-order derivates, followed by bandpass frequency filtering of the BOLD timeseries between 0.008 Hz and 0.09 Hz.

The scans were then parcellated into resting-state networks using the Oxford-Harvard atlas^25–27^, which is commonly used in neuroimaging software, and hence provides a widely accepted reference for brain region parcellation. In additional analyses, the resulting networks from the Independent Component Analysis (ICA) and the templates from the Power’s atlas^28^ were used to compare the results. We were interested in seven main resting-state networks: the DMN, sensorimotor, visual, salience, dorsoattentional, frontoparietal (executive), and language networks (Fig 1).

**Figure 1.**
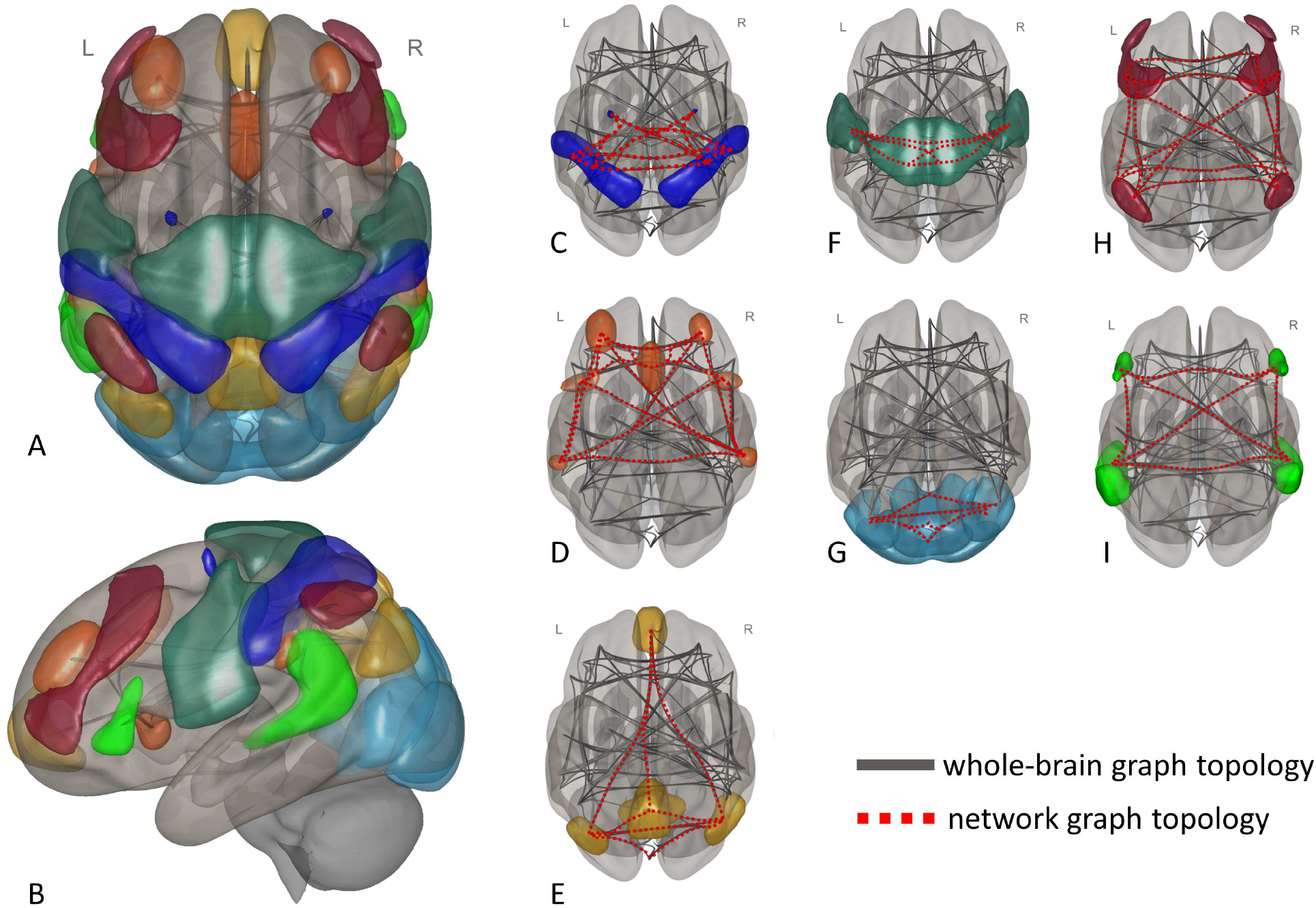
Resting-state networks from the Oxford-Harvard. A) The top-down view of the spatial maps of all seven networks on the 3D brain. B) The lateral view of the spatial maps of all seven networks on the 3D brain, C) The graph topology of the Dorsoattentional network, D) The graph topology of the Frontoparietal network, E) The graph topology of the Default Mode network, F) The graph topology of the Sensorimotor network, G) The graph topology of the Visual network, H) The graph topology of the Salience network, I) The graph topology of the Language network.

### 2.3. First- and second-level analysis

Regions of interest connectivity matrices were estimated characterising the patterns of functional connectivity. Functional connectivity strength was represented by Fisher-transformed bivariate correlation coefficients from a weighted general linear model (GLM), defined separately for each pair of target areas, modelling the association between their BOLD signal timeseries. To compensate for possible transient magnetisation effects at the beginning of each run, individual scans were weighted by a step function convolved with an SPM canonical haemodynamic response function and rectified.

Group-level analyses were performed using a GLM. For each individual voxel a separate GLM was estimated, first-level connectivity measures at this connection as dependent variables, and groups as independent variables. Inferences were performed at the level of individual clusters, based on parametric statistics from Gaussian Random Field theory. Results were thresholded using a combination of p<0.05 connection level threshold, a cluster-forming p<0.001 voxel-level threshold, and a corrected p-FDR <0.05 cluster-size threshold. The data were subsequently entered into a graph theory analysis.

### 2.4. Graph theoretical analysis

Graphs are data structures that model pairwise relations between network components. When applied to fMRI data, ‘nodes’ represent different brain regions and ‘edges’ represent connections between them.

Subsets of regions of interest were selected for each network using available regions from the Oxford-Harvard atlas, which were then analysed in the second-level FDR-corrected within-network analyses with groups as factors (*APOEe4* carriers vs non-carriers), and age, gender, and years of education as covariates. A binary connectivity matrix is obtained by thresholding the values of the correlation matrix. This step is crucial to eliminate spurious connections in the data; however, the choice of the threshold is often arbitrary, and the different methods of thresholding may lead to substantial variability in the data^29^. Since we were interested in retaining the strongest connections, we utilised the cost method with the threshold of 0.15 to keep the 15% of the strongest connections. In additional analyses, we explored alternative thresholding by using the z-scores.

Seven graph theoretical properties were explored to assess the topological patterns of different networks, namely the measures of centrality (i.e., Betweenness Centrality, Closeness Centrality, Degree Centrality) and the measures of network structure and efficiency (i.e., Average Path Length, Global Efficiency, Clustering Coefficient). A detailed description of mathematical calculations and roles of each property can be found elsewhere^30,31^ and is summarised in Table 1.

**Table 1.**
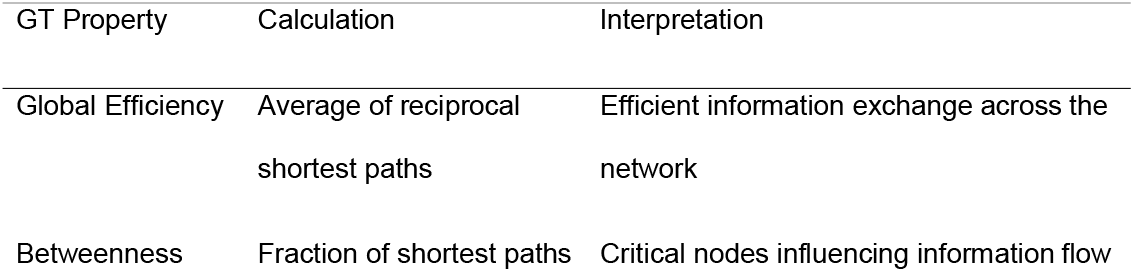
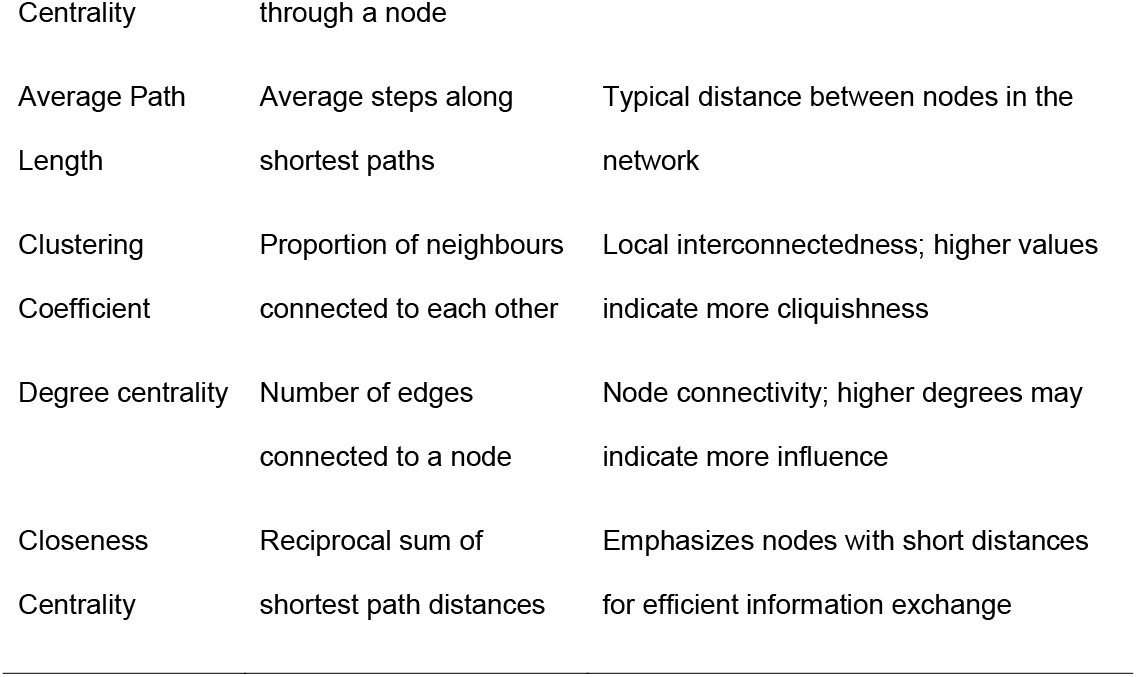
The summary of the Graph Theory properties studied in the present article.

Briefly, Betweenness Centrality measures the number of the shortest paths running through a node, identifying critical hubs for efficient information flow. Closeness Centrality assesses how closely connected a node is to others, pinpointing key locations essential for efficient information transfer. Average Path Length of a network is calculated as the average number of steps along the shortest paths for all possible pairs of nodes in the network, so it reflects the typical distance between pairs of nodes in the network. Global efficiency averages the inverses of the shortest path lengths between all pairs of nodes in the network, representing effective communication across the entire network. Clustering coefficient measures which nodes in the network tend to cluster together, indicating local interconnectedness. Degree Centrality refers to the number of connections per node and thus reflects the importance of a node in the network.

## 3. Results

### 3.1. Whole-brain connectivity differences

To assess the baseline whole-brain connectivity differences between *APOEe4* carriers and non-carriers, the Oxford-Harvard atlas was used to parcellate the brain into 48 cortical and 21 subcortical regions. The graph theoretical analysis of all regions was then conducted using the same methods as for the analysis of the networks. There were no significant differences found between the *APOEe4* carriers and non-carriers on any of the graph theoretical measures (p-FDR>0.05).

### 3.2. Graph Theory differences between the *APOEe4* carriers and non-carriers

Graph Theory analysis revealed differences in functional connectivity between young *APOEe4* carriers and non-carriers in four networks in the Average Path Length and Closeness Centrality properties (Table 2).

**Table 2.**
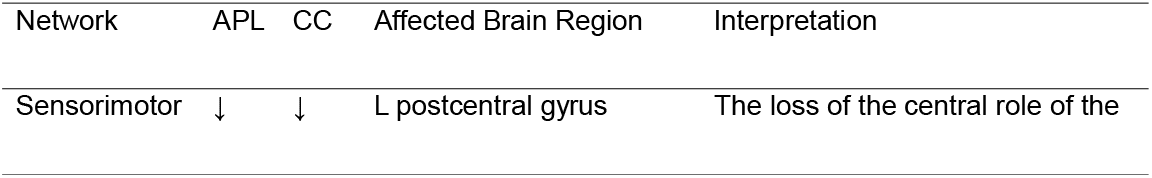
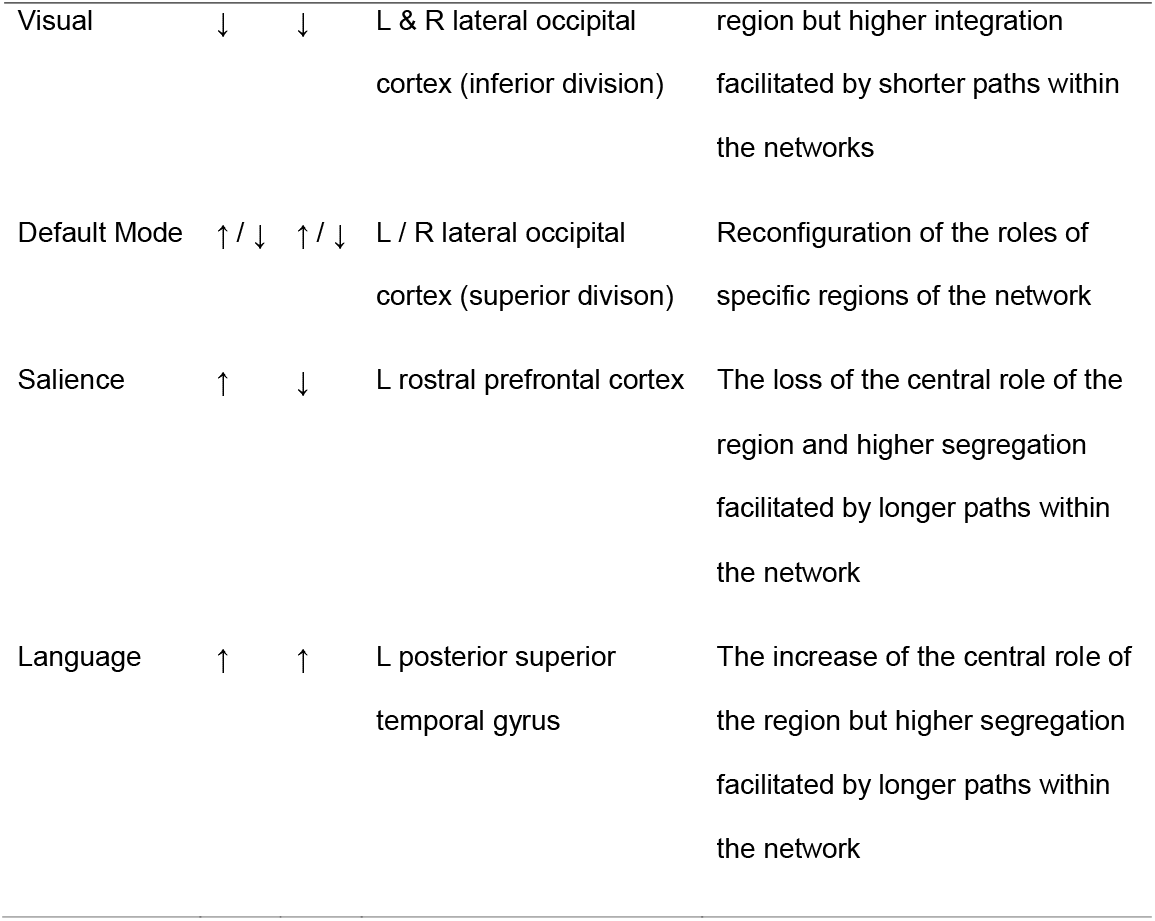
The summary of the significant results in Average Path Length (APL) and Closeness Centrality (CC) in *e4* carriers relative to non-carriers and their interpretation in the context.

Average Path Length and Closeness Centrality decreased in *APOEe4* carriers in the left postcentral gyrus in the sensorimotor network (−55, -12, 29; T = -5.64, p-FDR < 0.0001; Fig 2A) and bilaterally in the inferior division of the lateral occipital cortex in the visual network (−37, -79, 10; 38, -72, 13; T = -5.23, p-FDR < 0.0001; Fig 2B). There were no significant differences in the sensorimotor network globally; however, Degree Centrality, Cost, and Global Efficiency showed a trend when using an uncorrected p-value (T = 2.02, p-unc = 0.0451). A similar trend was observed in Cost and Global Efficiency in the visual network (T = 2.02, p-unc = 0.0451).

**Figure 2.**
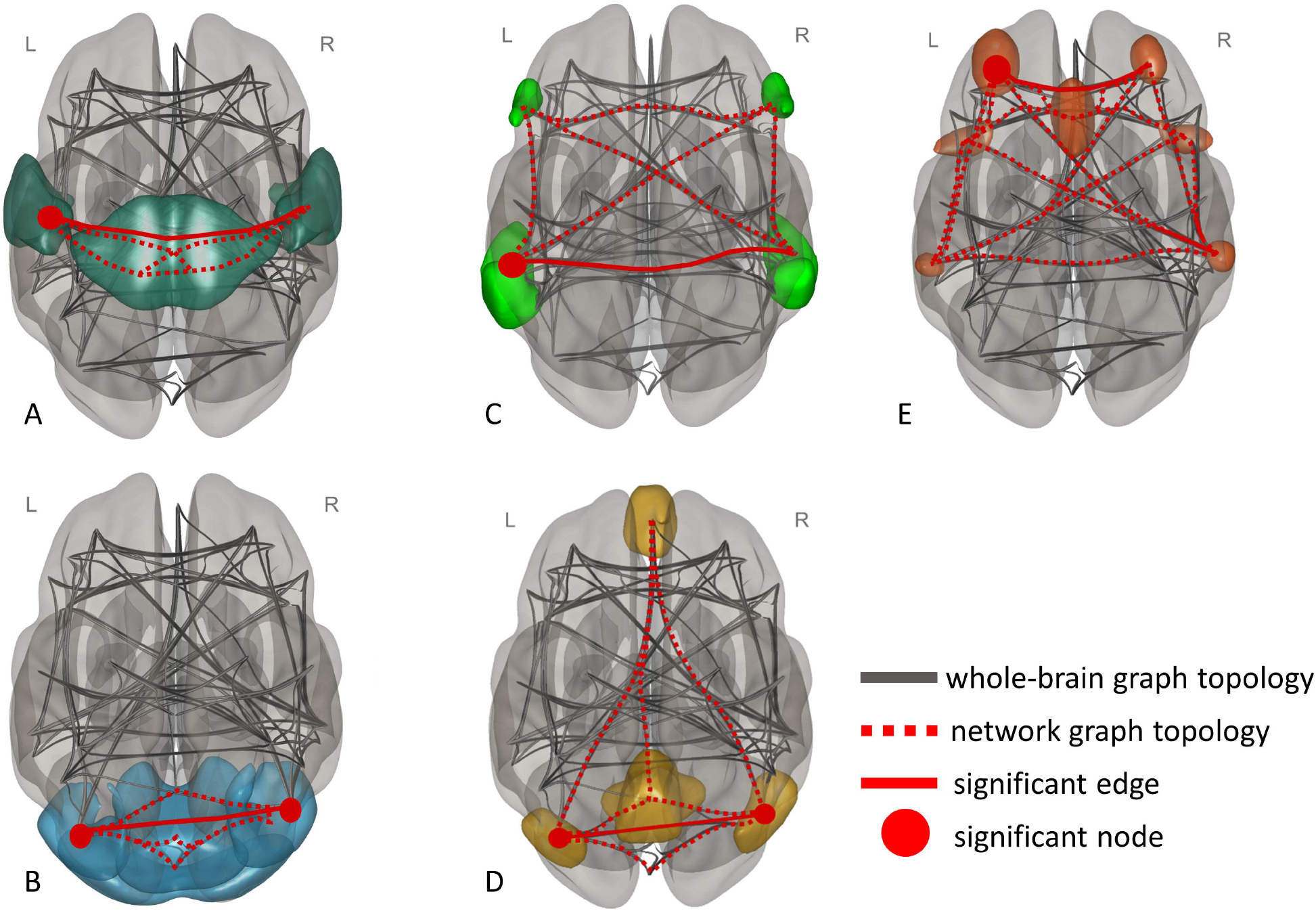
The visual representation of the topology of graphs including significant between-group differences in resting-state networks and respective brain nodes. The topology of the graph is represented by dotted lines, the significant edges are represented by solid lines, and the significant hubs are represented by circles. A) Sensorimotor network (left postcentral gyrus). B) Visual network (bilateral inferior division of the lateral occipital cortex). C) Language network (left posterior superior temporal gyrus) D) DMN (bilateral superior division of the lateral occipital cortex). E) Salience network (left rostral prefrontal cortex).

The opposite pattern was found in the language network, where Average Path Length and Closeness Centrality increased in the left posterior superior temporal gyrus in *e4* carriers (−57, -47, 15; T = 2.71, p-FDR = 0.0316; Fig 2C).

The DMN and salience network showed interesting patterns of differences in graph-theoretical properties. Firstly, a mixed directionality was observed in the DMN. While Average Path Length and Closeness Centrality decreased in *APOEe4* carriers in the right superior division of the lateral occipital cortex (47, -67, 33; T = -2.81, p-FDR = 0.0117; Fig 2D), they increased in the left superior division of the lateral occipital cortex (−39, -77, 33; T = 4.74, p-FDR < 0.0001; Fig 2C). The DMN did not show any global differences on any of the properties; however, an uncorrected trend was observed in Cost and Global Efficiency (T = 2.02, p-unc = 0.0451). There was a simultaneous increase in Average Path Length (−32, 45, 27; T = 2.98, p-FDR = 0.0261; Fig 2E) and a decrease in Closeness Centrality in the left rostral prefrontal cortex in the salience network (−32, 45, 27; T = -2.78, p-FDR = 0.0459; Fig 2D). Similarly, no global differences in networks were observed; yet, almost all graph theoretical properties showed trends when using the uncorrected values, including Global Efficiency (T = 2.17, p-unc = 0.0322), Betweenness Centrality (T = 2.02, p-unc = 0.0455), Closeness Centrality (T = 2.03, p-unc = 0.0446), Cost (T = 1.99, p-unc = 0.0484), Average Path Length (T = 2.01, p-unc = 0.0461), and Degree Centrality (T = 2.01, p-unc = 0.0464). Similar global trends were observed across graph theory properties in the remaining networks; yet, no results reached the level of significance when controlling for multiple comparisons.

When comparing the results to the results derived using z-scores as a thresholding method with a correspondingly set threshold level, we observed full or partial consistency in the sensorimotor, visual, language, and salience networks, which highlights the internal validity of the findings from these networks. Our findings demonstrate the sensitivity to different brain parcellation methods. The detailed results of the replications by using different thresholding methods and different brain parcellation methods derived from the ICA and Power’s atlas are summarised in Supplementary Materials 1.

## 4. Discussion

In this study, we investigated the graph-theoretical properties of the resting-state networks in young adults with *APOEe4* allele. The systematic literature review of brain structure and function in young *APOEe4* carriers revealed no previous studies have utilised graph theory to study networks in this demographic^23^; hence, to the best of our knowledge, this is the first study to explore the properties of multiple resting-state networks of *APOEe4* carriers in young adulthood. We found significant differences in functional connectivity in the DMN, salience, language, sensorimotor, and visual networks. Interpreting whether differences in graph-theoretical properties are either beneficial or detrimental is challenging due to the inherent interdependence among network characteristics, regional roles, and broader contextual factors. To comprehensively assess the significance of observed changes, those need to be carefully considered and are discussed below.

### 4.1. The sensorimotor and visual networks

A change in Average Path Length implies the average distance between pairs of nodes in the networks has shortened or prolonged. A simultaneous change in Closeness Centrality of a specific region indicates a shift in its role within the network and is expected due to the mathematical relationship of these properties. When there is a simultaneous decrease in these properties, the region is losing its centrality in terms of efficient communication with the broader network, but it is also becoming more locally interconnected. In other words, the information exchange between the nodes in the network is facilitated by shorter paths, contributing to a more tightly integrated and responsive network.

This pattern was observed in the left postcentral gyrus in the sensorimotor network and bilaterally in the inferior division of the lateral occipital cortex in the visual network. Speculatively, this could imply that these regions, while less influential in terms of global network dynamics, are fostering more efficient and direct communication within their surroundings. A decrease in Path Length was previously observed in a whole-brain network in AD^32^. This could be interpreted as the disruption of the ‘small-world behaviour’ which was also observed in other studies in AD^33^ and in cognitively healthy adults with AD-related pathology^34^, albeit by using different mechanisms. In contrast with our study, Sanz-Arigita and colleagues explored whole-brain connectivity in symptomatic AD. The analysis of the whole brain did not yield any significant differences between *APOEe4* carriers and non-carriers in our study.

Rewiring of the sensorimotor network, which is involved in the integration of sensory information with motor commands, has been observed in MCI and AD^35^ and seems to be modulated by the *APOE* genotype^36^. Similar findings were reported in visual networks in task-based studies in both MCI and AD^37^, indicating its association with functions such as spatial localisation, face recognition, or motor perception that are also progressively disturbed in AD. However, the evidence from cognitively healthy individuals is still limited.

### 4.2. The DMN, salience, and language networks

Both the DMN and the salience network showed a more complex pattern of mixed directionality. While the Average Path Length and the Closeness Centrality decreased in the right superior division of the lateral occipital cortex, they increased in the left superior division of the lateral occipital cortex, which might suggest that the DMN is reconfiguring the communication patterns and roles of these specific regions. The regions of the parieto-occipital cortex have been previously linked with early AD pathology such as tau accumulation^38^. The authors proposed the relationship between the posterior DMN connectivity and tau burden. The underlying mechanisms are still not clear; however, the disturbances in functional connectivity in the DMN, specifically its posterior regions, have been consistently reported and studied extensively in MCI and AD (reviewed^39^) and healthy middle-aged adults with genetic risk factors, mainly the *APOEe4* allele (reviewed^6^).

There was an increase in Average Path Length and a decrease in Closeness Centrality in the left rostral prefrontal cortex in the salience network. Unlike the pattern seen in the sensorimotor and visual networks, this opposing directionality points to a decrease in network efficiency. The left rostral prefrontal cortex may be becoming less central in terms of efficient communication with the broader network and the typical distance required to travel between the regions connecting the left rostral prefrontal cortex is increasing. This region has been previously linked with network efficiency mediated by education in elderly individuals with a high proportion of *APOEe4* carriers^40^. Moreover, broader areas of the prefrontal cortex show consistent disturbances in brain function and structure in middle-aged *APOEe4* carriers (reviewed^5,41^). Both DMN and salience networks play crucial roles in multiple cognitive processes; therefore, differences in topology in their functional connectome may signify the alterations in fundamental neural processes that could contribute to susceptibility to neurodegenerative diseases later in life.

A simultaneous increase in Average Path Length and Closeness Centrality that was observed in the language network of *APOEe4* carriers might suggest a consolidation of the network’s structure. The left posterior superior temporal gyrus seems to be establishing longer paths to reach other nodes in the language network, indicating greater centrality in facilitating information exchange across the network. However, this increase in centrality may come at the expense of the local interconnectedness, potentially leading to a more segregated network structure with fewer direct connections between directly neighbouring nodes. Segregation of functional networks is a common phenomenon in AD that has been associated with cognitive performance^42^ and resilience^43^, or tau pathology accumulation^44^.

### 4.3. The limitations and future directions

To account for the limited number of brain regions in the Oxford-Harvard atlas, we explored the networks derived from the Power’s atlas and the ICA. While our study demonstrated robustness across various graph-theory thresholding methods, the findings were sensitive to different parcellation methods. The choice of brain parcellation can significantly influence outcomes, as seen in the ageing population^45,46^, possibly due to the brain atlas concordance problem^47^ and might contribute to the observed heterogeneity in functional neuroimaging findings across young *APOEe4* carriers^23^. Currently lacking a standardised parcellation atlas in neurological and psychiatric research, future studies should carefully consider their choice, enhancing the interpretability, generalisability, and clinical applicability of neuroimaging findings. This consideration may also shed light on the mixed directionality of connectivity differences observed in young *APOEe4* carriers.

While functional connectivity has already contributed substantially to the research of AD, the focus has predominantly been on symptomatic populations and at-risk middle-aged individuals. Detecting changes in various networks, such as the DMN, years before the occurrence of clinical symptoms, and correlating the connectivity measures with cognitive improvement post-treatment^48^ indicates the potential of functional connectivity in pre-symptomatic research. However, introducing more sophisticated computational approaches, including mathematical and machine learning methods sensitive to subtle brain changes, can significantly enhance efforts to map vulnerability, even in early adulthood^16^. This can also increase our understanding of the underlying mechanisms of brain changes discussed thoroughly in our previous work^49^. By leveraging longitudinal and multimodal setups, researchers can establish trajectories of brain alterations associated with major AD risk factors, such as the *APOEe4* allele, and their impact on other brain and cognitive changes throughout the lifespan.

Variations in genetic markers, such as the *APOEe4* allele, may differently impact brain connectivity depending on ethnic background^14^. Therefore, it is crucial that studies properly account for and report ethnicity to ensure accurate interpretations of neuroimaging data. Our study adds to the limited body of evidence that *APOEe4* impacts the brain structure and function in young Chinese Han adults. Recognising these ethnic-specific factors can aid in generalising conclusions about brain health and disease susceptibility across different populations and should be of focus in future studies.

Investigating risk and resilience factors associated with AD throughout the lifespan is essential for biomarker development, early detection of disease, and uncovering novel targets for therapeutic intervention. However, early adulthood remains a significantly underexplored period. Yet, the findings may provide the necessary context for how early brain vulnerabilities occur, what mitigates them, and what other factors have a protective effect. For example, exploring the effect of the *e2* allele, which is associated with a lower risk of AD-related neurodegeneration, might uncover new targets for research on promoting life-long health^50^. The present study lacked the statistical power to delve into the effect of *e2* comprehensively, emphasising the need for future research to address this gap. Understanding both similarities and differences in the mechanisms via which *e4* and *e2* alleles affect the brain structure and function is imperative for developing strategies for AD prevention and therapeutic intervention.

## 5. Conclusions

Using graph theoretical analysis of resting-state fMRI, our study showed that in young cognitively healthy adults, Closeness Centrality and Average Path Length were consistently affected in the DMN, salience, sensorimotor and visual networks, in the brain areas that have been previously linked with structural, functional, or pathological changes in AD. Moreover, this study also underscores the critical role of the methodological choices, particularly in the selection of a brain parcellation method and the subsequent need for the detailed reporting of the methodological choices.

## 6. Data availability

The anonymised data from the present study are available from the corresponding author on reasonable request.

## Acknowledgements

LK conceptualised and designed the study, performed the first-level analysis, drafted the manuscript, and created figures and tables. LK and AB performed the second-level and graph theory analyses. JZ collected the data. CMN pre-processed the data. JZ, CRW, and LS set up the extension of the PREVENT-Dementia study. LS, CMN, GMT, CWR, and JOB provided the feedback. LS secured the funding, provided the guidance, and oversaw the study. We thank the members of the Artificial Intelligence and Computational Neuroscience at the University of Sheffield for their insights, comments, and meaningful discussions.

