## Supplementary documents for "Cognitively healthy young adults with *APOEe4* gene show disrupted functional connectivity of graph properties in multiple resting-state networks"

Table 1. The full summary of all significant between-group results (APOEe4 carriers vs non-carriers) across different types of brain parcellation and using different thresholding methods.

| Network | Thresholding method | Parcellation method | GT property | Significant results |
| --- | --- | --- | --- | --- |
| DMN | Cost | Oxford-Harvard | Closeness centrality | T=4.74, p-FDR=0.000027; T=-2.81, p-FDR=0.011792 |
| DMN | Cost | Oxford-Harvard | Average path length | T=4.74, p-FDR=0.000027; T=-2.81, p-FDR=0.011792 |
| DMN | Cost | Power’s | Degree | T=3.87, p-FDR=0.010148 |
| DMN | Z-score | Power’s | Degree | T=3.87, p-FDR=0.010149 |
| Salience | Cost | Oxford-Harvard | Closeness centrality | T=-2.78, p-FDR=0.045885 |
| Salience | Cost | Oxford-Harvard | Average path length | T=2.98, p-FDR=0.026170 |
| Salience | Z-scores | Oxford-Harvard | Global efficiency | T=-3.22, p-FDR=0.011302 |
| Salience | Z-scores | Oxford-Harvard | Degree | T=-3.09, p-FDR=0.017098 |
| Salience | Z-scores | ICA | Betweenness Centrality | T=2.99, p-FDR=0.040489 |
| Sensorimotor | Cost | Power’s | Average path length | T=3.23, p-FDR=0.049972 |
| Sensorimotor | Cost | Oxford-Harvard | Average path length | T=-5.64, p-FDR=0.000001 |
| Sensorimotor | Cost | Oxford-Harvard | Closeness centrality | T=-5.64, p-FDR=0.000001 |
| Sensorimotor | Z-scores | Oxford-Harvard | Average path length | T=-3.14, p-FDR=0.002120 |
| Sensorimotor | Z-scores | Oxford-Harvard | Closeness centrality | T=-3.14, p-FDR=0.002120 |
| Visual | Cost | Oxford-Harvard | Average path length | T=-5.23, p-FDR=0.000001 |
| Visual | Cost | Oxford-Harvard | Closeness centrality | T=-5.23, p-FDR=0.000001 |
| Visual | Z-scores | Oxford-Harvard | Average path length | T=-5.23, p-FDR=0.000002 |
| Visual | Z-scores | Oxford-Harvard | Closeness centrality | T=-5.23, p-FDR=0.000002 |
| Frontoparietal | Cost | Power’s | Closeness centrality | T=3.22, p-FDR=0.022690 |
| Frontoparietal | Z-scores | Oxford-Harvard | Closeness centrality | T=-2.61, p-FDR=0.042393 |
| Frontoparietal | Z-scores | Oxford-Harvard | Average path length | T=2.61, p-FDR=0.042393 |
| Dorsoattentional | Cost | ICA | Closeness centrality | T=-3.64, p-FDR=0.001235 |
| Dorsoattentional | Z-scores | ICA | Closeness centrality | T=-2.68, p-FDR=0.025484 |
| Dorsoattentional | Z-scores | ICA | Average path length | T=2.68, p-FDR=0.025484 |
| Dorsoattentional | Z-scores | ICA | Degree | T=3.40, p-FDR=0.003672; T=2.74, p-FDR=0.009585; T=2.73, p-FDR=0.009585 |
| Dorsoattentional | Z-scores | ICA | Global efficiency | T=3.28, p-FDR=0.004885;  T=3.06, p-FDR=0.004885;  T=2.93, p-FDR=0.004885;  T=2.87, p-FDR=0.004885 |
| Dorsoattentional | Z-scores | ICA | Betweenness centrality | T=2.78, p-FDR=0.012525 |
| Language | Cost | Oxford-Harvard | Average path length | T=2.71, p-FDR=0.031670 |
| Language | Cost | Oxford-Harvard | Average path length | T=2.71, p-FDR=0.031670 |
| Language | Z-scores | ICA | Betweenness centrality | T=2.79, p-FDR=0.049165 |
